# Psychometric Validation of a Clinician-Reported Clinical Severity Assessment in *STXBP1-Related Disorder*

**DOI:** 10.64898/2026.05.27.26354243

**Authors:** Megan Abbott, Peter Jacoby, Katie Angione, Tim A. Benke, Hsiao-Tuan Chao, JoeyLynn Coyne, Kristin Cunningham, Danielle deCampo, Jenny Downs, Jim Goss, Zachary Grinspan, Morgan Jolliffe, Juliet Knowles, Eric Marsh, Jillian L. McKee, Andrea Miele, Samuel R. Pierce, Sarah M. Ruggiero, Charlene Son Rigby, Megan Stringfellow, Sarah Tefft, Katherine Xiong, Ingo Helbig, Scott Demarest

## Abstract

**AIM:** STXBP1-related disorder (STXBP1-RD) is a severe developmental and epileptic encephalopathy characterized by early-onset seizures and persistent cognitive and motor impairments. With disease-modifying trials emerging, a disorder-specific severity scale is needed. To address this, we adapted a validated clinician-reported measure from CDKL5 Deficiency Disorder to develop the STXBP1 Clinical Severity Assessment (S-CSA) and evaluated its psychometric properties.

**METHOD:** The S-CSA was adapted from the CDKL5 Clinical Severity Assessment through expert consensus sessions with STXBP1 clinicians. Revisions addressed gaps in motor and vision domains, adding tremor and vision items. The measure was administered to 123 individuals with STXBP1-RD. Psychometric evaluation included confirmatory factor analysis, internal consistency, composite reliability, average variance extracted, and distinctiveness, compared with recommended thresholds.

**RESULTS:** Analyses supported a three-domain structure (motor, communication, vision) with factor loadings >0.5 and strong internal consistency (Cronbach’s alpha >0.7; composite reliability >0.88). Model fit and variance metrics met recommended standards, and domains demonstrated distinctiveness. No ceiling or floor effects were observed. Minimal skew was seen in motor (0.34) and communication (−0.16) domains; positive skew in vision (2.2) was seen, identifying patients with and without cortical visual impairment.

**INTERPRETATION:** The S-CSA demonstrates strong validity and reliability in *STXBP1*-RD and may show utility in clinical trials for *STXBP1*-RD and potentially other severe DEEs.

*What this paper adds:*

- First disorder-specific clinician-reported severity scale developed for *STXBP1*-RD.
- Demonstrated strong reliability and validity across three core domains.
- S-CSA has now shown content and psychometric validity.
- S-CSA may be a useful endpoint for upcoming *STXBP1* clinical trials.

## 1. Introduction

*STXBP1*-related disorder (*STXBP1*-RD) is a severe developmental and epileptic encephalopathy (DEE) characterized by early-onset seizures and developmental delay, caused by variants in the *STXBP1* gene. *STXBP1*-RD is one of the most common genetic causes of DEE, with a prevalence of 3.8 per 100,000 births.^1, 2^ Epilepsy is but one facet of the disorder with many other components including intellectual disability, hypotonia, cortical visual impairment, and movement disorder.^3^ A disease concept model for *STXBP1*-RD identified multiple symptoms beyond seizures that are important to patient families including developmental delay, behavior, expressive communication and motor issues.^4^ These patient family priorities were corroborated in the *STXBP1-RD* Externally-led Patient Focused Drug Development Meeting held with the FDA in October 2023.^5^ There are currently no comprehensive outcome measures that capture these important features (communication, tremor, cortical visual impairment) of *STXBP1*-RD. Several disease modifying therapies (DMTs) are currently in various stages of development for *STXBP1-*RD. However, the lack of accurate clinical outcome measures that capture meaningful change is emerging as a major issue in the field. *STXBP1*-RD is a complex neurodevelopmental disorder with a wide range of clinical features reported in the literature. While improvement in seizures is often considered an intuitive endpoint for DEEs, emerging data suggests that the seizure trajectory in *STXBP1*-RD is too volatile to serve as a primary endpoint.^6, 7^ Consequently, DMT trials in *STXBP1*-RD are at risk of failing if focused on seizure improvement alone.^8^ *STXBP1*-RD would benefit from alternative endpoints apart from seizure frequency.

Recent efforts to improve clinical trial readiness for *CDKL5* Deficiency Disorder (CDD) may be relevant for *STXBP1*-RD, given the similarities in features between the two DEE conditions. The *CDKL5* Clinical Severity Assessment (CCSA) was introduced in 2019 following a modified Delphi approach to address the need for a multidimensional severity measure for CDD.^9^ Because clinicians and caregivers contribute distinct types of information, the instrument was divided into two independent components, each of which underwent formal content validation using cognitive interviewing techniques.^10, 11^ The clinician-focused component, known as the CCSA-Clinician, is designed to capture disease features that can be directly evaluated during routine clinical encounters. It is a relatively quick measurement compared to other scales (takes clinician 15-30 minutes to complete). Validation of this measure involved eight think-aloud interviews with neurologists experienced in CDD, followed by a structured consensus review.^10^ The finalized clinician instrument includes 27 items encompassing neurological deficits, functional status, and associated comorbid conditions.^10^ The CCSA-Clinician then underwent a psychometric validation process through which three domains or “factors” were identified: vision, communication and motor skills. Children with neurodevelopmental disorders often present with complex clinical features and understanding the interplay between vision, communication, and motor development can be difficult. Despite these challenges, the specific items in the CCSA-Clinician performed and mapped well to the three domains, all of which had sufficiently high factor loadings and no clear floor or ceiling effects.^10^

The aim of this study was to evaluate the psychometric validity of the CCSA Clinician in *STXBP1*-RD. *STXBP1-RD* and CDD share many similarities as severe DEEs. Both conditions include severe, drug-resistant epilepsy, developmental delay, comorbid medical complexities (such as poor feeding and respiratory complications), cortical visual impairment, and significant behavioral challenges.^4, 6, 12^ However, subtle differences between the two disorders exist, requiring adaptation of the CCSA for *STXBP1*-RD prior to piloting. This study presents the processes of adaptation followed by piloting of the *STXBP1*-Clinical Severity Assessment (S-CSA) through the natural history study to determine the psychometric validity and reliability in this new population.

## 2. Methods

### 2.1 Population

The STARR (*STXBP1* Clinical Trial Readiness) study was utilized for this work. This is a multicenter, prospective natural history study that conducts in-person visits with individuals with *STXBP1*-RD every six months, enabling the piloting and longitudinal collection of S-CSA data in this population. Participants from both the Children’s Hospital of Philadelphia (CHOP) and Children’s Hospital Colorado (CHCO) sites were recruited. Data were collected and managed using Research Electronic Data Capture (REDCap) electronic data capture tool.^13, 14^ This study was approved by the Internal Review Board at the University of Pennsylvania IRB 23-021140 and University of Colorado COMIRB 24-0647 with patient consent given for the conducting of this research.

### 2.2 Consensus sessions and measure modification

Two consensus sessions were held with a group of *STXBP1* clinical experts involved in the STARR study to ensure the group felt the measure adequately covered the *STXBP1* phenotype. This discussion were informed by the STXBP1 Disease Concept Model.^4^ While the measure was largely felt to be comprehensive for *STXBP1*, assessment of tremor and more subtle presentations of visual impairment were determined to be gaps in the current CCSA. Three tremor items and three additional vision items were drafted and edited by the group in subsequent sessions until all parties were satisfied with the final S-CSA. The S-CSA was then performed on all participants seen at CHCO and CHOP for the STARR study. Assessors were trained using standardized training manuals and instructional videos that have been developed to support consistent administration of the measure.

### 2.3 Measure

The final S-CSA following these consensus sessions and modeled off the CCSA-Clinician^10^, is an electronic clinician questionnaire completed via REDCap immediately following the administration of a structured clinical examination. Administration of the S-CSA takes approximately 15-30 minutes. The S-CSA consists of 35 items, organized in 3 domains: (motor-18 items, communication-8 items, vision-9 items).

The number of gross motor items administered varies by the child’s age. Items assessing head control and transitions from supine to sitting are administered across all age groups, while additional items assessing sitting (≥7 months), sit-to-stand transitions (≥15 months), standing (≥15 months), and walking (>18 months) are measured based on the child’s age. A similar graduated item system based on age is applied to the communication domain. Scores for the three domains (motor, communication, and vision) are calculated by averaging the scores of the items administered. Each item is converted to a 100-point scale for calculation of domain scores and the total score is the average of the domain scores, with higher scores indicating greater severity.

### 2.4 Statistical Analysis

T-tests (for continuous variables) and chi-square tests (for categorical variables) were conducted to compare demographic characteristics and total S-CSA scores between the two sites, assessing for significant differences between the two cohorts.

Confirmatory factor analysis (CFA) was conducted on the S-CSA using the previously validated CCSA domain structure identified for *CDKL5* Deficiency Disorders, including vision, communication, and motor domains. Sample size calculation: Our target number of participants is guided by the rigorous sample size requirements of factor analysis. The recommended sample size for a Confirmatory Factor Analysis (CFA) to ensure precision of parameter estimates, lack of bias and solution propriety can be anywhere between 30 and 460 depending on the number of factors, the number of items per factor and the size of factor loadings, with an overall rule of thumb for 3 factors being a minimum sample size of 100.^15^ Duplicate participant records resulting from visits at both sites were removed by using a common study ID, retaining only the first site visit. The weighted least squares (WLS) estimator appropriate for ordinal items was used. Factor loadings were computed for each item. Model fit was evaluated using the Root Mean Square Error of Approximation (RMSEA) and Tucker-Lewis Index (TLI), and these values were compared against predefined, widely accepted thresholds.^16^ Where necessary, items were removed or reallocated across factors to achieve satisfactory model fit. Internal consistency for each domain was assessed with Cronbach’s alpha, and the Average Variance Extracted (AVE) was calculated for each domain in the final CFA models. Distinctiveness of the domains was evaluated using the Fornell-Larcker criterion, which compares each domain’s AVE with the square of the maximum inter-domain correlation.^17^ Domain scores (average of item scores) and a total score (average of domain scores) for the S-CSA were calculated and the distribution of domain and total scores assessed. Ceiling and floor effects were defined as having >10% of the cohort with the minimum (0s) or maximum scores (100s).

## 3. Results

### S-CSA

There were 123 S-CSA questionnaires from STARR participants with no missing data. Demographic characteristics of the participants are shown in Table 1. There were no statistically significant differences (p-value <0.05) between the 2 sites for demographics or overall S-CSA score. The motor and vision domain scores were also not significantly different. The communication score did differ significantly between the sites with Children’s Hospital of Philadelphia having higher (more severe) communication scores.

**Table 1:** Demographic Information Across Cohort.

| Characteristic | Children's Hospital Colorado (n=47) | Children's Hospital of Philadelphia (n=76) | Total (n=123) | P-Values (* for <0.05) |
| --- | --- | --- | --- | --- |
| **Median Age, (IQR) in years** | 8.5, (3.9-12.7) | 6.3, (3.1-11.2) | 7.7, (3.4-12, 0.85) | 0.85 |
| **Age range: n (%)** |  |  |  |  |
| 0-5 years | 15 (32%) | 29 (38%) | 44 (36%) |  |
| 5-10 years | 12 (25%) | 23 (30%) | 35 (28%) |  |
| 10-15 years | 14 (30%) | 9 (12%) | 23 (19%) |  |
| 15+years | 6 (13%) | 15 (20%) | 21 (17%) |  |
| **Gender: n (%)** |  |  |  |  |
| Male | 19 (40%) | 42 (56%) | 61 (50%) | 0.11 |
| Female | 28 (60%) | 34 (44%) | 62 (50%) |  |
| **CCSA Score** |  |  |  |  |
| Total Score Median (IQR) | 30.3 (20.3-39.2) | 31.8 (29.8-54.3) | 31.1 (22.8-43.2) | 0.06 |
| Motor Median (IQR) | 28 (13-47.2) | 28.75 (20.4-50) | 28.3 (17-50) | 0.37 |
| Communication Median (IQR) | 40 (31.7-59.6) | 60 (40-69.1) | 52.5 (35-63.8) | 0.001* |
| Vision Median (IQR) | 11.1 (5-25) | 6.7 (5-18.3) | 10 (5-23) | 0.67 |

A 3-factor model vision, communication, and motor domains was confirmed. This model continued the prior framework of combining the gross and fine motor function items with items describing muscle tone to form a Motor and Movement domain, the communication and alertness items to form a Communication domain and a Vision domain with items relating to fixing and following, observation of optokinetic nystagmus (OKN Drum; https://apps.apple.com/cz/app/okndrum-optokinetic-drum/id1438597800), eye alignment, roving and nystagmus. Importantly, the new tremor and vision items had to be removed to achieve adequate fit. The three tremor items tested included duration, distribution, and impact on gross motor function. Although tremor was present in 54% of participants, only 19% reported that it contributed to gross/fine motor disabilities. The 3-factor model displayed adequate fit with acceptably high (>0.5) factor loadings. The factor structure and factor loadings are shown in Table 2.

**Table 2:** Results of Psychometric Analyses.

| Summary of Psychometric Analyses & Accepted Thresholds |  |  |  |  |  |
| --- | --- | --- | --- | --- | --- |
| Analysis Type | Description | Statistical Measure and Threshold<sup>18</sup> | S-CSA Validation values |  |  |
|  |  |  | Motor | Communication | Vision |
| Factor Analysis | Statistical method that finds patterns in test responses by grouping together items that seem to be measuring the same idea into factors | Factor Loading (>0.5) | >0.78 | >0.66 | >0.55 |
|  |  | RMSEA (<0.08) | 0.095 |  |  |
|  |  | Tucker-Lewis Index (>0.9) | 0.975 |  |  |
| Internal Consistency | Indicates how well the questions on a test or survey all measure the same thing | Cronbach's Alpha (>0.7) | 0.852 | 0.894 | 0.797 |
|  |  | Composite Reliability (>0.7) | 0.958 | 0.880 | 0.865 |
| Avg. Variance Extracted | Shows how much of the information in a group of items reflects the concept being measured, compared to random error | Average Variance Extracted (AVE) (>0.5) | 0.820 | 0.648 | 0.576 |
| Distinctiveness | Evaluates that a concept being measured is clearly different from other concepts. | Max Correlation Squared (<AVE) | 0.517 | 0.517 | 0.349 |

For the 3-factor model, the Tucker-Lewis Index (TLI, 0.975) fit statistic was satisfactory. RMSEA was above the recommended upper threshold of 0.08 but literature suggests that the RMSEA is not recommended as a fit measure in models with few degrees of freedom and small sample size.^19^ Average Variance Extracted (AVE) for all domains (Range 0.58-0.82) was above the recommended minimum of 0.5. Cronbach’s alpha values for the domains ranged from 0.88 to 0.96 indicating satisfactory internal consistency. The maximum inter-domain correlation squared was less than AVE for all three domains (Communication, Vision, Motor), indicating that the Fornell-Larcker criterion for distinctiveness was satisfied for all domains (See Table 2)^17^.

Figures 1 and 2 illustrate the distributions of scores for each of the S-CSA domain scores and for the total score. For the total score, motor domain, and communication domain no floor or ceiling effects were present. The vision score shows a ceiling effect, with 15% of the cohort scoring the minimum (least severe) score but no floor effect. Minimal skew was seen in the overall score and communication/motor domains. Significant right skew was seen in the vision domain.

**Figure 1:**
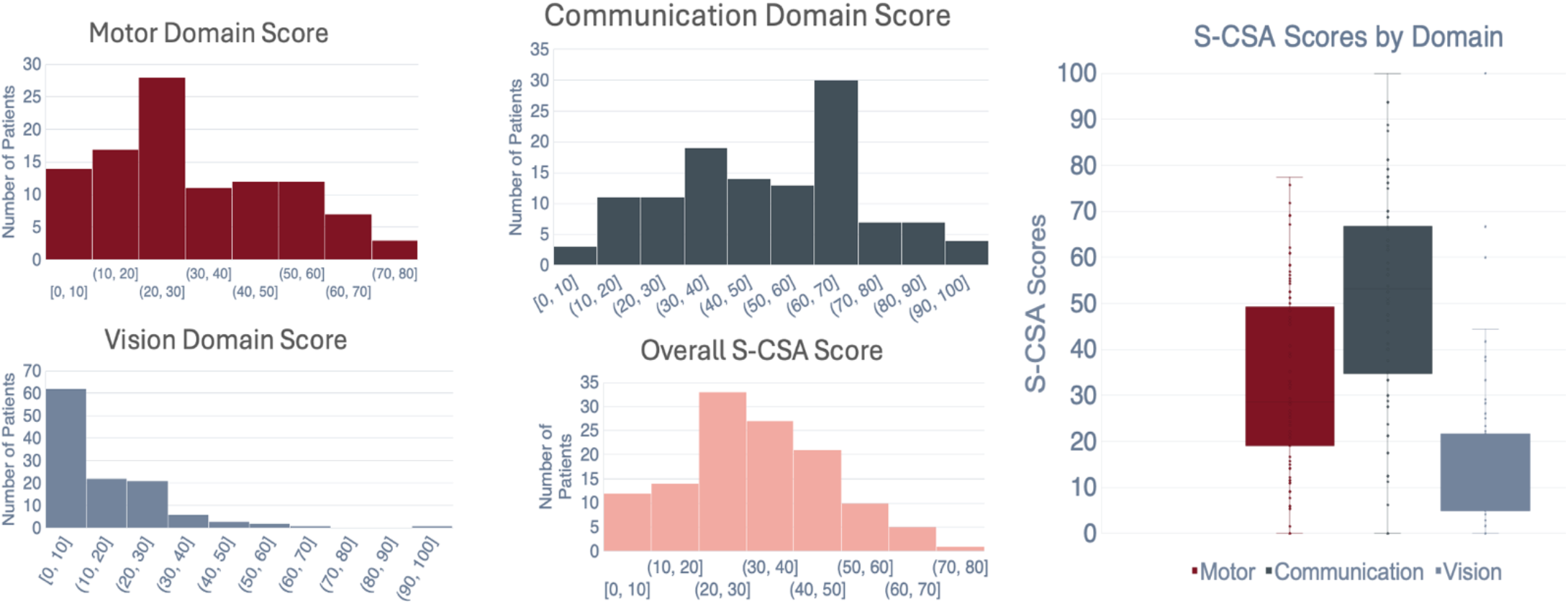
Distributions and rage of each domain score and the overall S-CSA.

**Figure 2:**
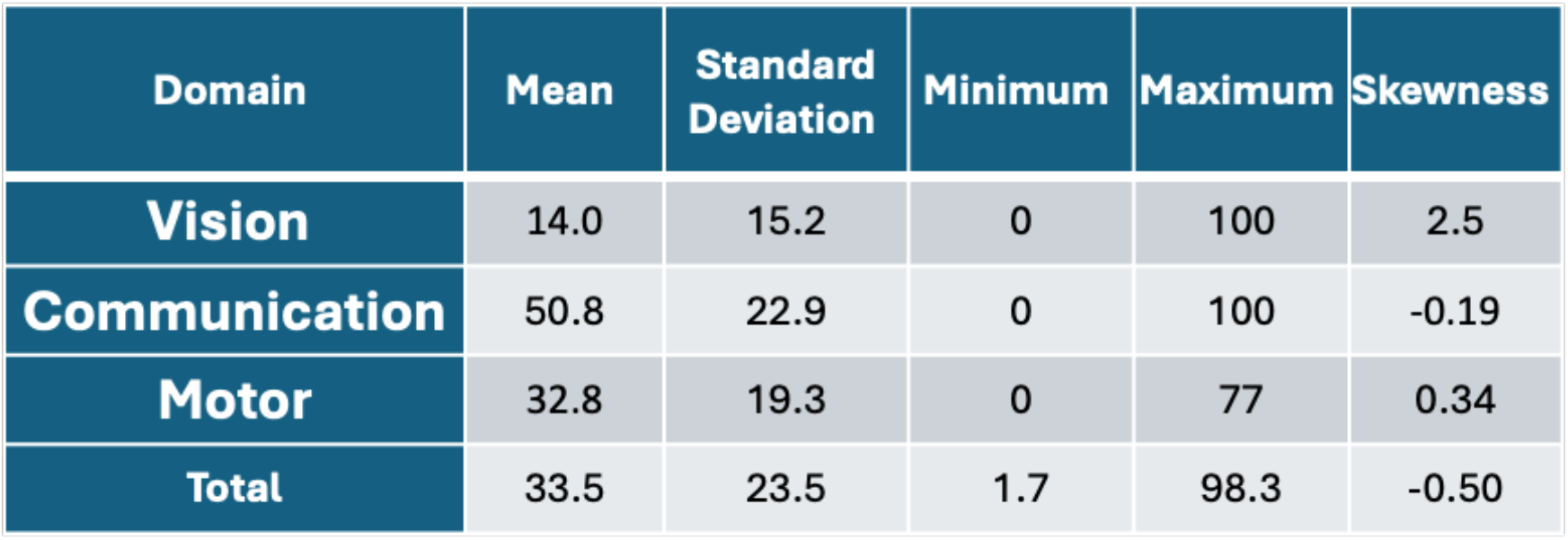
S-CSA Descriptive Statistics.

## 4. Discussion

The S-CSA is now a confirmed valid form of severity assessment for individuals with *STXBP1*-related disorder. Through confirmatory factor analysis, we demonstrated that the model fit was strong in all domains that were previously identified for a different DEE, highlighting important similarities between both conditions. Importantly, no floor or ceiling effects were observed in the overall score, a key component for measures intended for natural history studies and clinical trials.

The factor analysis led to the removal of the newly added Tremor and Vision items due to their poor fit within a coherent factor structure. The need to remove these items to have the key three domains emerge parallels our prior experience in *CDKL5* Deficiency Disorder. During the initial factor analysis of the original CCSA, several items related to movements, behavior, and autonomic function needed be excluded. For both *CDKL5* Deficiency Disorder and *STXBP1*-RD, potential reasons for item exclusion include scoring subjectivity, unpredictability within participants (tremor only present at certain times of day), a lack of correlation with clinical severity, or limited variability within the sample. Although the intention had been to expand the measure with additional tremor and vision items, our main theory is that these did not meaningfully contribute to overall clinical severity across the cohort, though they might for an individual patient, and therefore did not align with the current factor structure. To support this, clinicians only reported tremor contributing to disability a minority of the time. This outcome illustrates an important and expected aspect of measure development: while clinicians may aim to comprehensively capture the full phenotype of a condition, not all features necessarily contribute substantially to disease severity across the population. Similar to processes carried out with the initial CCSA in *CDKL5*, excluded items in the S-CSA for *STXBP1*-RD could still be important when considering natural history/clinical phenotyping, but may not be useful when measuring severity in *STXBP1*-RD. Therefore, we recommend collecting data for all items for descriptive purposes, with responses recorded in a non-scored “additional items” section at the end of the S-CSA.

The assessment of the vision domain on S-CSA for *STXBP1* showed important differences to the CCSA in *CDKL5*. Both assessments used the same items, but the distribution of vision domain scores was vastly different. Whereas vision domain scores in *CDKL5* were roughly normally distributed, we observed a prominent right skew in S-CSA (*STXBP1)* vision domain scores. The skew was towards the lower scores, indicating fewer symptoms. The differences in the distribution of visual domain scores is likely explained by the differences in the two genes’ phenotypes. *CDKL5* has a cortical visual impairment incidence of greater than 75%, while *STXBP1-*RD only has a cortical visual impairment incidence of 50%; in addition, when CVI is present in *STXBP1*-RD, it tends to be milder.^12, 20^ Therefore the skew towards less impairment in the visual domain in participants with *STXBP1-RD* highlights a unique and unexpected strength of the S-CSA: it not only provides a psychometrically robust measure of *STXBP1*-RD severity, but also distinguishes differences in visual impairment severity between two separate conditions. This skew did not affect the overall domain score, and we believe accurately reflects the range of visual impairment seen in patients with *STXBP1*-RD.

The general lack of floor or ceiling effects suggests the measure captures the full breadth of severity seen in *STXBP1*-RD. The measures of reliability, internal consistency and discriminant validity were all within accepted thresholds. These robust psychometric measurements provide evidence that the S-CSA is a valid measurement of severity in this population and support its use in future clinical trials for *STXBP1-RD*. The initial CCSA-Clinician for CDD, and now the S-CSA for *STXBP1*-RD, takes an innovative approach of quantifying neurologic examination findings. This tactic allows clinician-judged severity to be measured consistently in a more granular manner than clinician global impression scales, which are commonly used in current clinical trials.

The strong performance of a minimally modified S-CSA in *STXBP1-RD* indicates potential applicability across other severe DEEs, which will be explored in future work. Development of multi-domain, robustly validated measures that accurately capture the range of severity in the DEE population could support broader clinical trial readiness. Therefore, the validation of the S-CSA in *STXBP1*-RD is a critical proof-of-concept that such measures can easily adapted to other DEEs and validated for use in future trials.

## 5. Limitations

Performance of the measure was completed at two sites, and though training was completed for the measure at both sites, variability in how the measure was administered could still exist between the two sites. There was a difference seen in the communication domain, which could either represent the Children’s Hospital of Philadelphia cohort having higher communication impairment or challenges with inter-rater reliability on the communication items. This will need to be investigated further with longitudinal assessment and inter-rater reliability measurements. That said, no statistical differences were seen between the two groups in terms of participant demographics or total S-CSA score. 123 participants, though adequate, is also a relatively small participant cohort for performing this type of psychometric analysis and if additional items were desired for testing in additional DEEs, a larger cohort of individuals would be required. The measure still needs to undergo longitudinal assessment to evaluate stability over time and test-retest reliability.

## 6. Conclusions

The S-CSA has evidence for reliability and validity in the *STXBP1-*RD population. This measure represents a novel way of measuring disease severity through clinician report and holds promise as an outcome measure for use in future clinical trials.

## Data Availability

All data produced in the present study are available upon reasonable request to the authors

## 7. Acknowledgements

We express our deepest gratitude to the *STXBP1-*RD patients, caregivers, and families in the community for their contributions, including the donation of their invaluable time. Their support was necessary to generate the data essential to this research.

We appreciate the contributions of the additional STARR investigators at the Baylor College of Medicine and Texas Children’s Hospital site: including Alvina Zia, Dr. Ekaterina Sanchez-Romero, Dr. Elaine Seto, Dr. Danielle S. Takacs, Dr. Sruthi Thomas, Roberta Olivares, Natasha Feuerbach, Dr. Kristen S. Fisher, Dr. Mikael Guzman-Karlsson, Arden Wheeler, and Dr. Andres Jimenez-Gomez; at the Children’s Hospital Colorado site: including Andrea Gerk, Megan Stringfellow, Brittany Gladfelter, Emily Wilson, Morgan Joliffe, Ann Reynolds, Kiara Hamlin, Julie Vanek, Kellie Sitarz, Kaitlyn Kennedy, and Dana Bennick; at the Stanford Medicine Children’s Health site: including Rayann Solidum, Sweta Patnaik, Amy Weisman, Sophia Magana, and Lauren Mattas; and at the Weill-Cornell Medicine site: including Dara Jones, Jennifer Cross, Carlianne Ward, Millie Stone, Natalie Wayland, and Aida Osis.

